# Dense Optic Nerve Head Deformation Estimated using CNN as a Structural Biomarker of Glaucoma Progression

**DOI:** 10.1101/2021.09.08.21263299

**Authors:** Ali Salehi, Madhusudhanan Balasubramanian

## Abstract

**Purpose:** To present a new structural biomarker for detecting glaucoma progression based on structural transformation of the optic nerve head (ONH) region.

**Methods:** A dense ONH deformation was estimated using deep learning methods namely DDCNet-Multires, FlowNet2, and FlowNet-Correlation, and legacy computational methods namely the topographic change analysis (TCA) and proper orthogonal decomposition (POD) methods using longitudinal confocal scans of the ONH for each study eye. A candidate structural biomarker of glaucoma progression in a study eye was estimated as average magnitude of flow velocities within the ONH region. The biomarker was evaluated using longitudinal confocal scans of 12 laser-treated and 12 contralateral normal eyes of 12 primates from the LSU Experimental Glaucoma Study (LEGS); and 36 progressing eyes and 21 longitudinal normal eyes from the UCSD Diagnostic Innovations in Glaucoma Study (DIGS). Area under the ROC curves (AUC) was used to assess the diagnostic accuracy of the candidate biomarker.

**Results:** AUROC (95% CI) for LEGS were: 0.83 (0.79, 0.88) for DDCNet-Multires; 0.83 (0.78, 0.88) for FlowNet2; 0.83 (0.78, 0.88) for FlowNet-Correlation; 0.94 (0.91, 0.97) for POD; and 0.86 (0.82, 0.91) for TCA methods. For DIGS: 0.89 (0.80, 0.97) for DDCNet-Multires; 0.82 (0.71, 0.93) for FlowNet2; 0.93 (0.86, 0.99) for FlowNet-Correlation; 0.86 (0.76, 0.96) for POD; and 0.86 (0.77, 0.95) for TCA methods. Lower diagnostic accuracy of the learning-based methods for LEG study eyes were due to image alignment errors in confocal sequences.

**Conclusion:** Deep learning methods trained to estimate generic deformation were able to detect ONH deformation from confocal images and provided a higher diagnostic accuracy when compared to the classical optical flow and legacy biomarkers of glaucoma progression. Because it is difficult to validate the estimates of dense ONH deformation in clinical population, our validation using ONH sequences under controlled experimental conditions confirms the diagnostic accuracy of the biomarkers observed in the clinical population. Performance of these deep learning methods can be further improved by fine-tuning these networks using longitudinal ONH sequences instead of training the network to be a general-purpose deformation estimator.

## INTRODUCTION

In progressive glaucoma, the optic nerve head (ONH) region undergoes structural changes and deformation including death of retinal ganglion cells^1^, changes in the architecture and mechanical properties of the collagenous lamina cribrosa^2^, and progressive loss of retinal nerve fibers^3^. These progressive ONH changes observable using optical imaging instruments can be estimated from longitudinal sequences of ONH exams using computational methods and form a basis of structural biomarkers for detecting glaucoma progression in clinics^4–6^. In this work, we present deep-learning models to estimate a dense / pixel-level structural deformation of the ONH from optical exams and a candidate structural biomarker based on the estimated ONH deformation for detecting glaucoma progression.

Optical flow of a 2D or 3D image sequence represents a 2D or 3D field of apparent motion of constitutive elements of the structure or a scene being imaged. Specifically, each pixel in the optical flow field captures the magnitude and direction of structural deformation. Previously, optical flow methods have been used for autonomous vehicle navigation^7,8^, non-contact strain estimation^9^, deformation of microscopic structures^10^ and several biomedical applications such as measuring strain distributions in bovine articular cartilage^11^, quantifying motion of orbital tissues for diagnosing orbital disorders^12^, measuring strain in the bovine retina^13^, deformation of the lamina cribrosa^14^, biomechanics of the knees^15^, quantifying heart wall motion^16^ and cardiac motion estimation^17^.

Recently, deep learning methods have been quite successful and surpassed other classical methods for solving pixel-level prediction problems^18^. FlowNet was the first convolutional neural network (CNN) developed for estimated optical flow from a pair of images of a dynamic scene^19^. Teney et al.^20^ developed a CNN inspired by the classical spatio-temporal motion-energy filters of Heeger et al^21^. In general, learning-based methods, without the need for computationally demanding optimization steps, are more efficient than many of the classical methods of optical flow estimation.

Supervised learning-based methods require thousands of retinal image sequences for learning to differentiate glaucomatous eyes from non-progressing and stable eyes^22^. Though glaucomatous conditions can be experimentally controlled using animal models and *in silico* models, it is impossible to know the true state of dense deformation that the ONH region undergoes due to elevated intraocular pressure levels.

In this work, we present a candidate structural biomarker of glaucoma progression derived from ONH deformation estimated using deep-learning based optical flow methods and compare their diagnostic accuracy with non-learning based biomarkers of glaucoma progression namely proper orthogonal decomposition method^4,5^ (POD) and topographic change analysis^6^ (TCA). One of the important advantages of these methods is that they can be trained with synthetic image sequences (non-retinal) with known deformation field. The biomarkers are validated using longitudinal confocal microscopy exams of primate eyes under controlled experimental glaucomatous conditions from the LSU experimental glaucoma study^23^ (LEGS) and of human eyes from the UCSD diagnostic innovations in glaucoma study^5^ (DIGS).

## METHODS

### Subjects

#### Primate Eyes from the LSU Experimental Glaucoma Study

We utilize a library of ONH topographies of 12 primates (24 eyes), built from the LEG study^23,24^ to evaluate the performance of the candidate biomarker under controlled experimental conditions. In brief, one eye of each primate was treated with laser to induce glaucoma (glaucoma induced study eye) and the other eye is untreated (contralateral normal eye). The ONH of both the glaucoma induced study eye and the contralateral normal eyes of all the primates were imaged every two weeks using TopSS confocal scanning laser ophthalmoscope at 10 field of imaging with a lateral digital resolution of 11^24^. During each imaging session, 6 ONH topographies of each eye were obtained. The IOP in the glaucoma-induced study eye of each of the 12 primates was elevated by treating the trabecular meshwork with argon laser. Baseline images were acquired before IOP elevation. All the follow-up images were aligned with pre-laser session images based on retinal vasculature before estimating ONH deformation from optical flow. Detailed description of the LEG study, TopSS confocal microscope and the image alignment procedure used are available elsewhere^23,24^.

A brief summary of the pre- and post-laser imaging sessions of all the primates is presented in Table 1.

**Table 1:** Summary of the LEG study imaging sessions used for assessing candidate biomarkers

| Subject ID | Number of imaging sessions |  |  |  |
| --- | --- | --- | --- | --- |
|  | Glaucoma induced eye |  | Contralateral normal eye |  |
|  | Pre-laser | Post-laser | Pre-laser | Post-laser |
| 1C | 3 | 6 | 3 | 6 |
| 1D | 3 | 10 | 3 | 13 |
| 1E | 3 | 22 | 3 | 22 |
| 1F | 3 | 16 | 3 | 16 |
| 1G | 3 | 10 | 3 | 10 |
| 1I | 3 | 22 | 3 | 22 |
| 1J | 3 | 16 | 3 | 15 |
| 1K | 3 | 10 | 3 | 10 |
| 1L | 3 | 4 | 3 | 6 |
| 1N | 3 | 4 | 3 | 4 |
| 1O | 3 | 10 | 3 | 13 |
| 1P | 3 | 12 | 3 | 14 |

#### Human Eyes from the UCSD Diagnostic Innovations in Glaucoma Study

Longitudinal HRT-II images from thirty six progressing eyes from 33 study participants and twenty one normal eyes from 20 study participants from the UCSD Diagnostic Innovations in Glaucoma Study (DIGS) were used for assessing the candidate biomarker in clinical population. A detailed description of these study eyes and longitudinal image sequences are available elsewhere^25^. In brief, glaucoma progression was defined based on optic disk changes from stereo-photo assessment or likely progression by Standard Automated Perimetry (SAP) guided progression analysis (GPA; Humphrey Field Analyzer, software ver. 4.2). In the normal eyes, there was no history of IOP *>* 22 and all the HRT scans were acquired within a short duration (median of 0.5 years). Demographic and HRT imaging details are presented in Table 2.

**Table 2:** Demographic summary of study participants from the UCSD Diagnostic Innovations in Glaucoma Study

|  | No. of Eyes<br>(No. of<br>Subjects) | Age in yrs:<br>Mean (95%<br>CI) | No. of HRT<br>Exams:<br>Median<br>(range) | Followup<br>in yrs:<br>Median<br>(range) |
| --- | --- | --- | --- | --- |
| **Progressing**<br>**Eyes** | 36 (33) | 64.7 (61.6,<br>67.7) | 5 (4 to 8) | 4.1 (2.4,<br>7.0) |
| **Longitudinal**<br>**Normal**<br>**Eyes** | 21 (20) | 57.4 (49.7,<br>65.1) | 4 (4 to 8) | 0.5 (0.2,<br>8.0) |

### Optical Flow Estimation Methods

A brief description of the modern learning-based optical flow estimation methods namely FlowNet and DDCNet-Multires used for detecting structural changes in the ONH are as follows. These methods are generic estimators of motion field from image sequences and specifically were trained to estimate structural deformation using generic (non-retinal) image sequences with known ground truth of the motion or deformation field.

#### FlowNet

*FlowNet* is the pioneer of supervised CNN-based models developed for learning optical flow tasks from ground truth motion vectors^26^. *FlowNet-Simple* is designed as an encoder-decoder structure. *Flownet-Correlation* is a variation of FlowNet-Simple that uses a custom layer called correlation layer to explicitly match feature maps extracted from each image in a sequence. Both methods lack the ability to recover high-resolution features needed to accurately estimate optical flow and clear motion boundaries. These methods use a variational approach as the last optional refinement to improve the estimation.

*Flownet2* improved upon the FlowNet by stacking multiple networks, adding a small-displacement estimation network, and by scheduling the order of presenting training data to the network^27^. In a stacked network, higher-level blocks take the estimated flows from the previous blocks alongside the copies of input images as their input. Some of these FlowNet-Simple blocks were trained on specific datasets to achieve higher accuracy in estimating smaller displacements. Their experiments show that using intermediate flow estimates to warp one of the images and using the difference between the warped image and the reference image further improves the performance of FlowNet2.

#### DDCNet-Multires: Network Architecture

DDCNets^28^ are a family of deep dilated convolutional neural networks whose architecture is based on the following three guiding principles: 1) to *preserve spatial and kinetic features* in the input image sequences throughout the network. This is accomplished by keeping extracted features within each layer of the network at their respective spatial order and avoiding spatial aggregation operations such as pooling. 2) To achieve a *large-enough receptive field* for the output neuronal units to enhance their ability to detect larger motions. Receptive field of a neuron in the output layer represents the spatial and temporal extent of visibility and access to neighboring locations in the input image sequences. Therefore, the effective receptive field of a network is a function of the network architectures i.e. arrangement and types of network layers as well as the learned network weights. And 3) to address the challenges in estimating motion field in the presence of heterogeneous motion dynamics along with large displacements, occlusion and scene homogeneity using the receptive field guided multiresolution analysis^29,30^.

In this work, we specifically investigate the utility of the DDCNet-Multires architecture for estimating optic nerve head deformation^31,30^. In DDCNets, a systematic use of dilated convolutional layers is used to achieve the desired spatial characteristics, shape and texture of the receptive field. In its present configuration, DDCNet-Multires has three cascaded DDCNet *sub-nets* namely one *flow feature extractor* sub-net and two *flow feature refiner* sub-nets with successively decreasing extents and characteristics of their effective receptive field. Each of these sub-nets generates intermediate flow estimates with successively improving resolution of the overall flow estimate (multiresolution strategy). In addition to these subnets, a *spatial feature extractor* module is used to generate illumination invariant feature sequences from the input image sequences and these invariant sequences are used by the DDCNet sub-nets for flow estimation. Figure 1, shows a detailed network architecture of the DDCNet-Multires. A detailed description of the DDCNet-Multires architecture and training details are also available elsewhere^30^.

**Figure 1:**
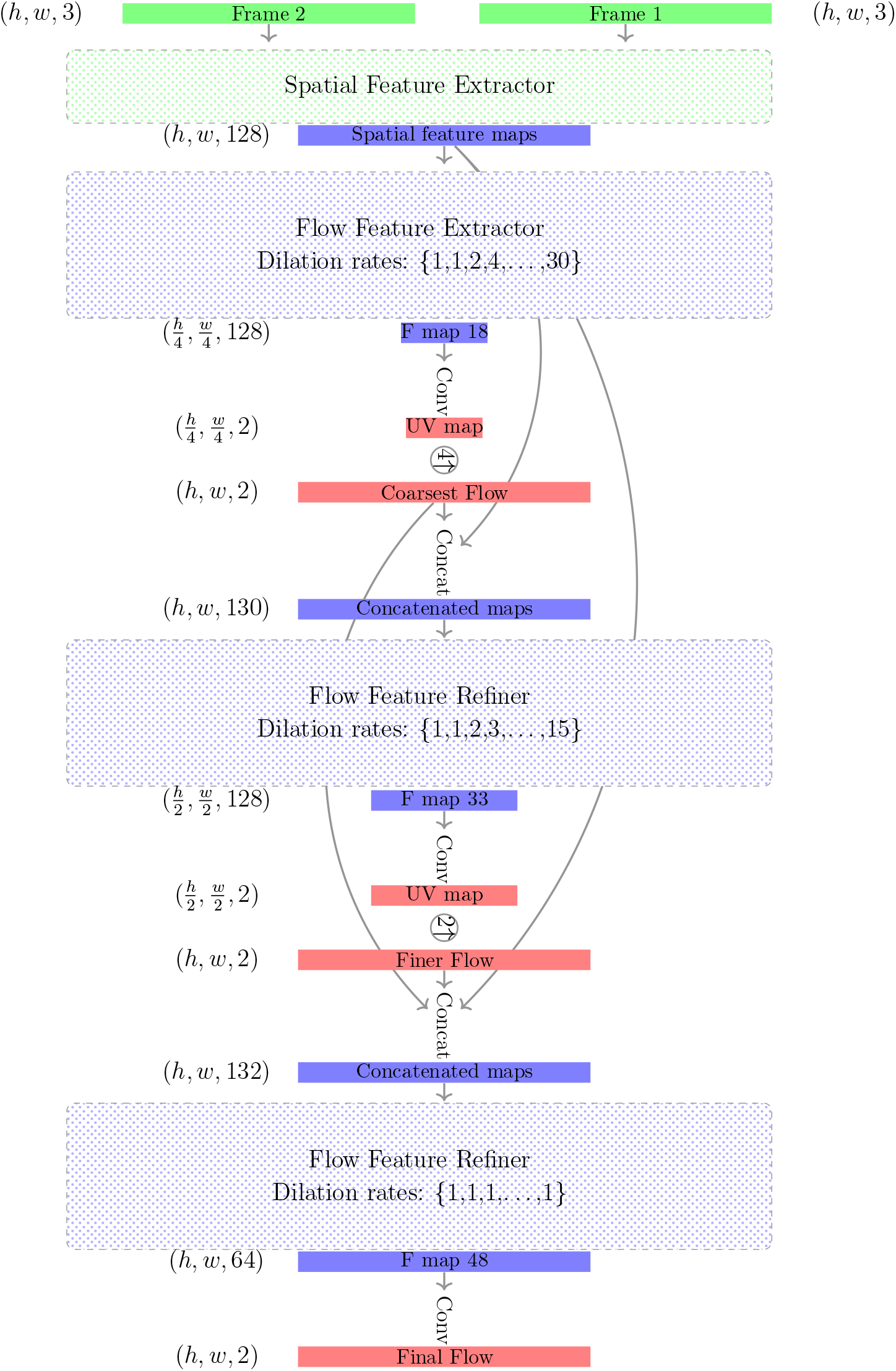
DDCNet-Multires: A multi-resolution network architecture built using one *spatial feature extractor* module, one *flow feature extractor* module, and two cascaded *flow feature refiner* modules.

#### DDCNet Multires: Training

The network was trained and tested using standard optical flow benchmark datasets namely the *Flying Chairs* dataset^19^, *Flying Things3D* dataset^32^, *MPI Sintel Dataset* ^33^, *KITTI2012* and *KITTI2015* datasets^34,35^ and *Middlebury* dataset^36^. A train-time augmentation method was used to improve the accuracy and generalization of the network to estimate ONH deformation^19^. The augmentation procedures included both geometric and photometric augmentations. It should be noted that the DDCNet-Multires training did not include any ocular images or ocular image sequences.

DDCNet-Multires was trained end-to-end using two graphical processing units (GPUs) with a batch size of 4 sequences and an initial learning rate of 0.0001 chosen heuristically. After 900k iteration, the learning rate was successively reduced by a factor of 2 when the network loss stayed flat for approximately 60k iterations. Network weights were initialized using the He-initialization method.

DDCNet-Multires trained on these generic image sequences was subsequently used for generating a quantitative measure of optic nerve head deformation from confocal image sequences of the optic nerve head.

### Candidate Structural Biomarker of Glaucoma Progression

For estimating ONH deformation at any given follow-up with respect to a baseline condition, reflectance images from the baseline and the follow-up exams were arranged in sequence and fed to DDCNet-Multires. For LEGS datasets, confocal reflectance images were not available and therefore topographic sequences were used. Using the DDCNet-Multires optical flow estimates, a measure of average flow magnitude within the optic disk region (candidate structural biomarker) was estimated for each of the ONH sequences. It is expected that the estimated average flow magnitude is lower for normal eyes when compared to eyes with glaucoma progression.

### Performance Analysis

Because the intraocular pressure of study eyes in LEGS study were experimentally controlled, ONH confocal scans from all post-laser treatment follow-up sessions were considered to be progressed and all the follow-up scans from the contralateral normal eyes were considered to be stable. Measures of candidate biomarkers estimated from each of the follow-up exams with respect to a pre-laser exam were used to assess their diagnostic performance in the LEGS study. Performance of these learning-based methods in this specific LEG study subject cohort was also compared with the classical TCA and POD computational approaches as reported in a previous study^24^.

In study eyes in DIGS study, measures of candidate biomarkers estimated between a baseline scan and the last follow-up scan were used to assess their diagnostic performance. Performance of these learning-based methods in this specific DIG study patient cohort was also compared with the classical TCA and POD computational approaches as reported in a previous study^5^.

The diagnostic performance of the candidate structural biomarker from each of the deep-learning methods was assessed using the area under their respective receiver operating characteristic curves (AUROC).

## RESULTS

Figure 2 shows the ROC curves of the candidate biomarkers from the learning-based methods in assessing contralateral normal eyes vs glaucoma-induced non-human primate eyes in the LEG study (Fig. 2a) and in assessing normal eyes vs glaucoma progressing eyes (human) in the DIG study (Fig. 2b). AUROC (95% CI) of the candidate biomarkers for the LEG study eyes were 0.83 (0.79, 0.88) for DDCNet-Multires; 0.83 (0.78, 0.88) for FlowNet2; 0.83 (0.78, 0.88) for FlowNet-Correlation; 0.94 (0.91, 0.97) for POD; and 0.86 (0.82, 0.91) for TCA methods. Among these methods, POD provided the highest diagnostic accuracy statistically significant from the learning-based methods (non-overlapping confidence interval measures of AUROC). For DIG study eyes, AUROC (95% CI) of the candidate biomarkers were 0.89 (0.80, 0.97) for DDCNet-Multires; 0.82 (0.71, 0.93) for FlowNet2; 0.93 (0.86, 0.99) for FlowNet-Correlation; 0.86 (0.76, 0.96) for POD; and 0.86 (0.77, 0.95) for TCA methods. FlowNet-Correlation provided the highest diagnostic accuracy statistically significant from the classical computational TCA and POD methods. Differences among the learning-based methods were not statistically significant (overlapping confidence interval measures of AUROC).

**Figure 2:**
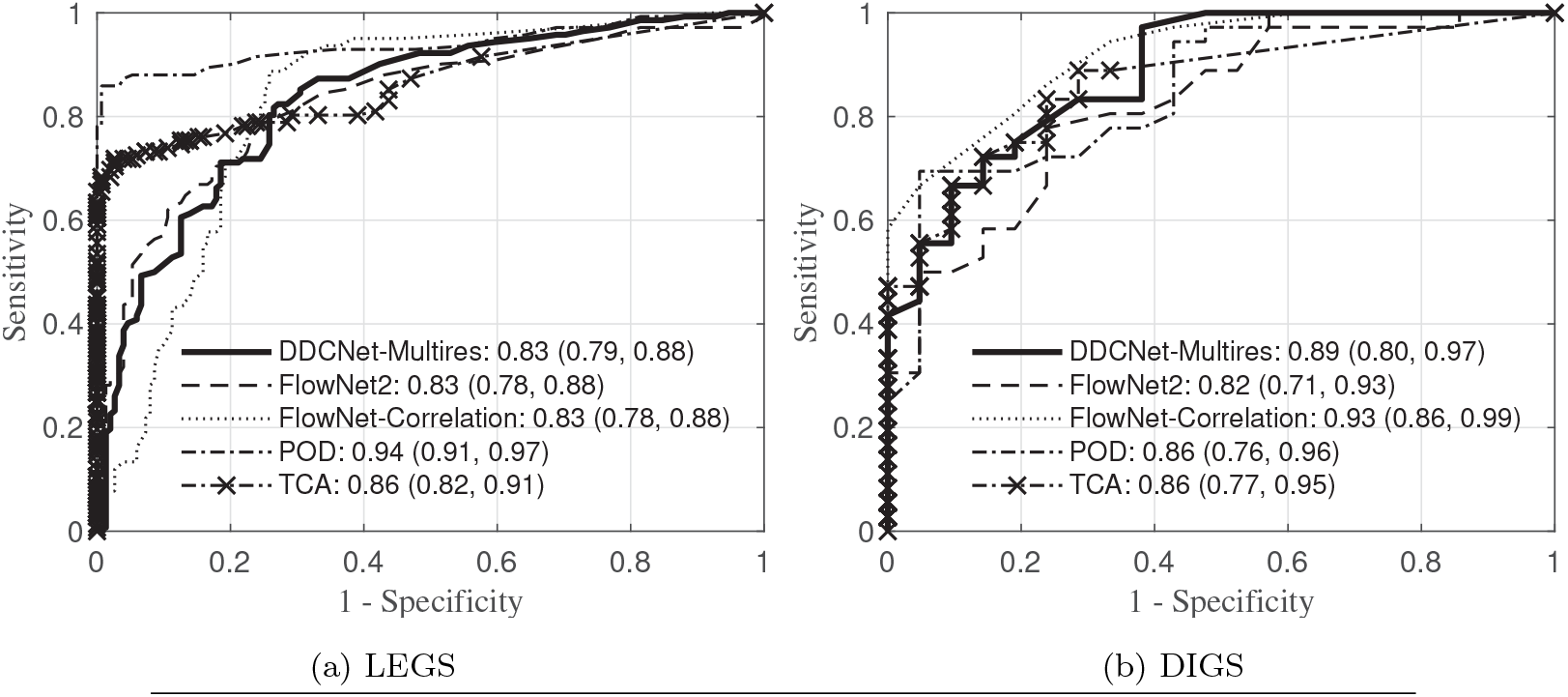
Receiver operating characteristics curves of learning-methods in detecting normal eyes and progressing eyes in the LEGS and DIGS datasets

Figure 4c shows the estimated deformation field in a contralateral normal eye (of subject 1D) in the LEG study estimated using a pre-laser confocal scan shown in Figure 4a and a post-treatment confocal scan shown in Figure 4b. The deformation field shown in Figure 4c is color-coded using a color-map shown in Figure 3 and represents the direction (color) and magnitude (color saturation) of deformation at each imaging location in the ONH scan. For visual inspection and inference, sparse motion vectors within the optic disk region (approximately for 5% of pixels) are also shown as an arrow or quiver plot. Similarly, Figure 5 shows an example of the ONH deformation field estimated in the corresponding glaucoma-induced study eye (subject 1D). A large deformation field estimated by the learning-based method for the glaucoma-induced eye (Figure 5c) agrees with the visually observable ONH damage in the longitudinal confocal scans (Figures 5a, Figure 5b). In the contralateral normal eye, the magnitude of the deformation estimated and visually observed from the confocal scans were smaller (Figure 4).

**Figure 3:**
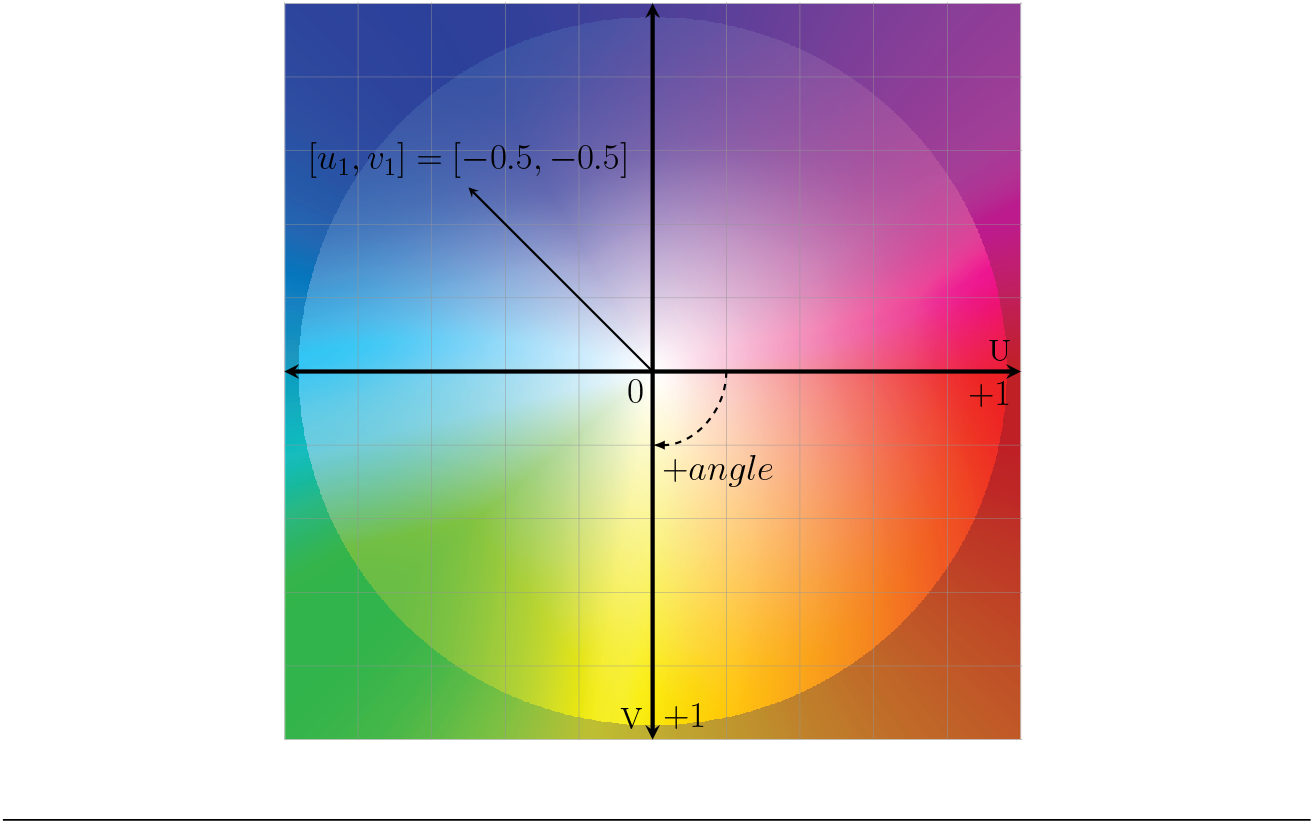
Color coding scheme generating flow map from motion vectors. The angle of motion at each pixel is represented by the color of the pixel and flow magnitude is visualized by the saturation of the pixel color. For example, flow vector [1, 0] is encoded as red and [−0.5, −0.5] is encoded as blue.

**Figure 4:**
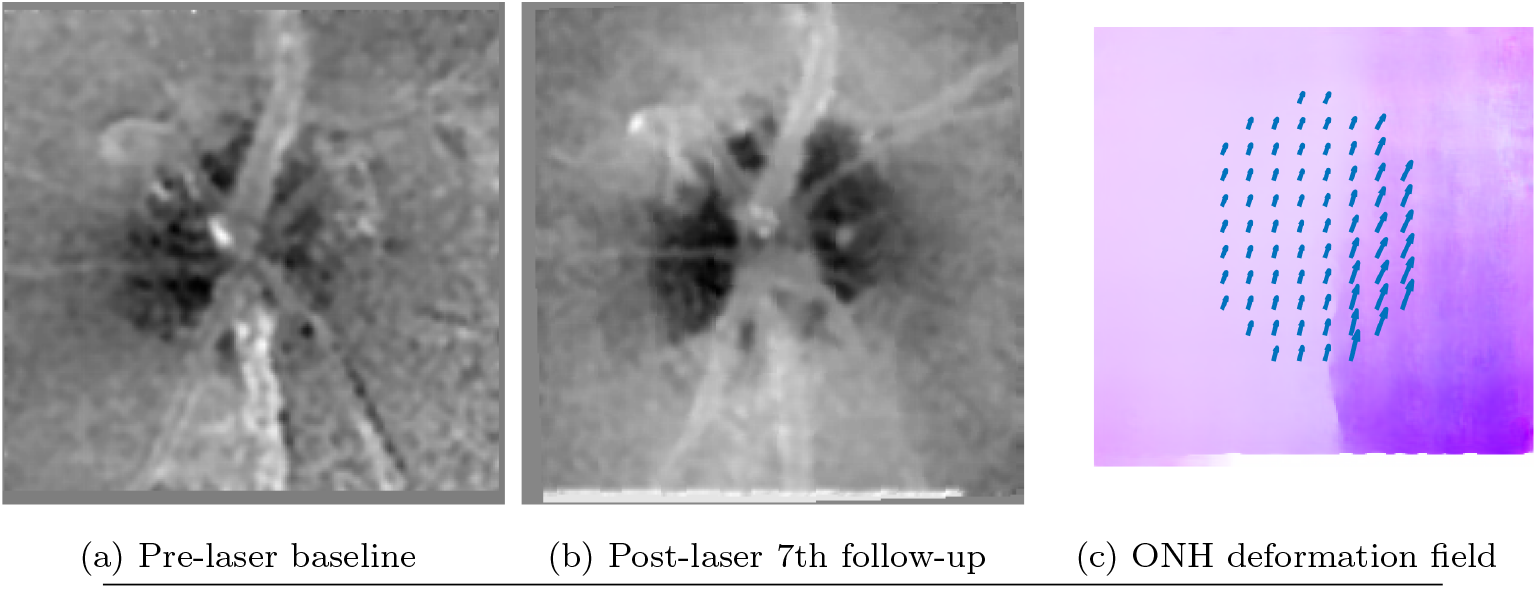
Example of ONH deformation observed in a LEGS contralateral normal eye of subject 1D (non-human primate). Deformation field estimated using the DDCNet-Multires method visualized using a color coding scheme shown in Figure 3. For quick inference, deformation field within the optic disk region also shown as arrow plot.

**Figure 5:**
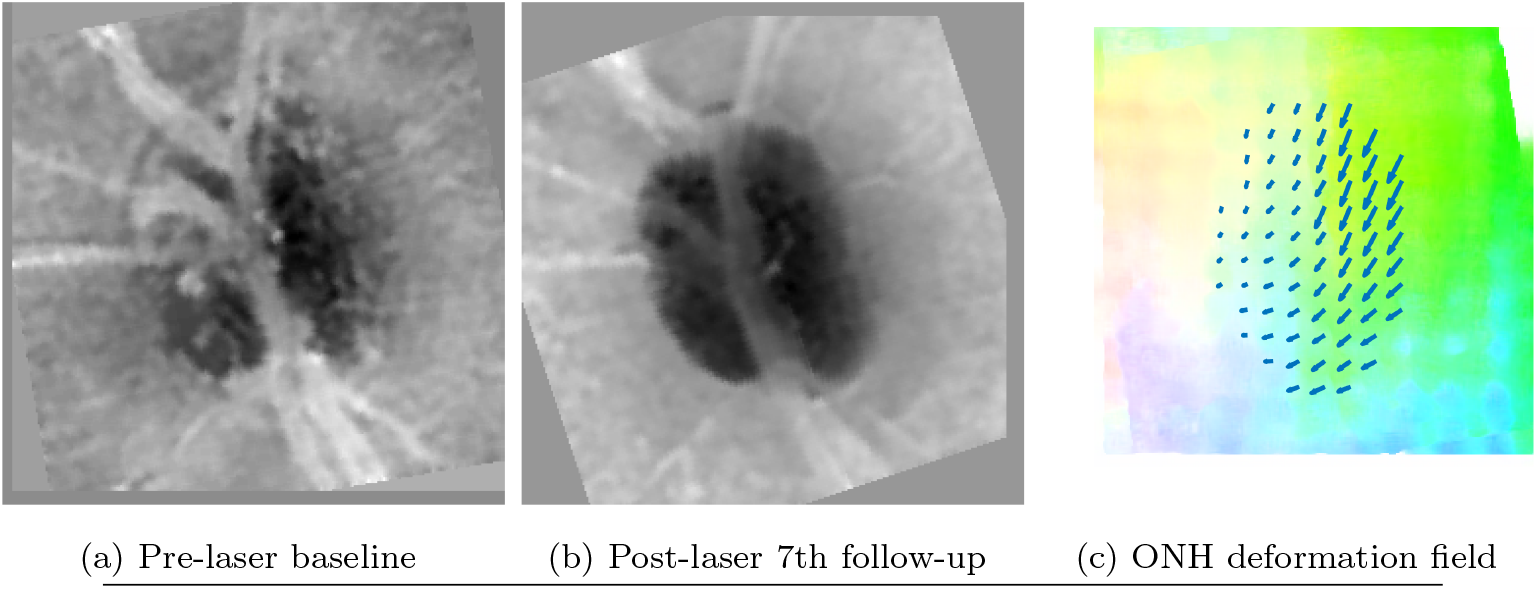
Example of ONH deformation observed in a LEGS glaucoma-induced study eye of subject 1D (non-human primate). Deformation field estimated using the DDCNet-Multires method visualized using a color coding scheme shown in Figure 3. For quick inference, deformation field within the optic disk region also shown as arrow plot.

Figures 6 and 7 show examples of deformation field estimated by the DDCNet-Multires method for a longitudinal normal eye and a glaucoma progressing eye respectively. As observed in the LEG study eyes, the magnitude of the deformation (higher color saturation in the deformation field) is higher in the progressing eye when compared to the normal eye.

**Figure 6:**
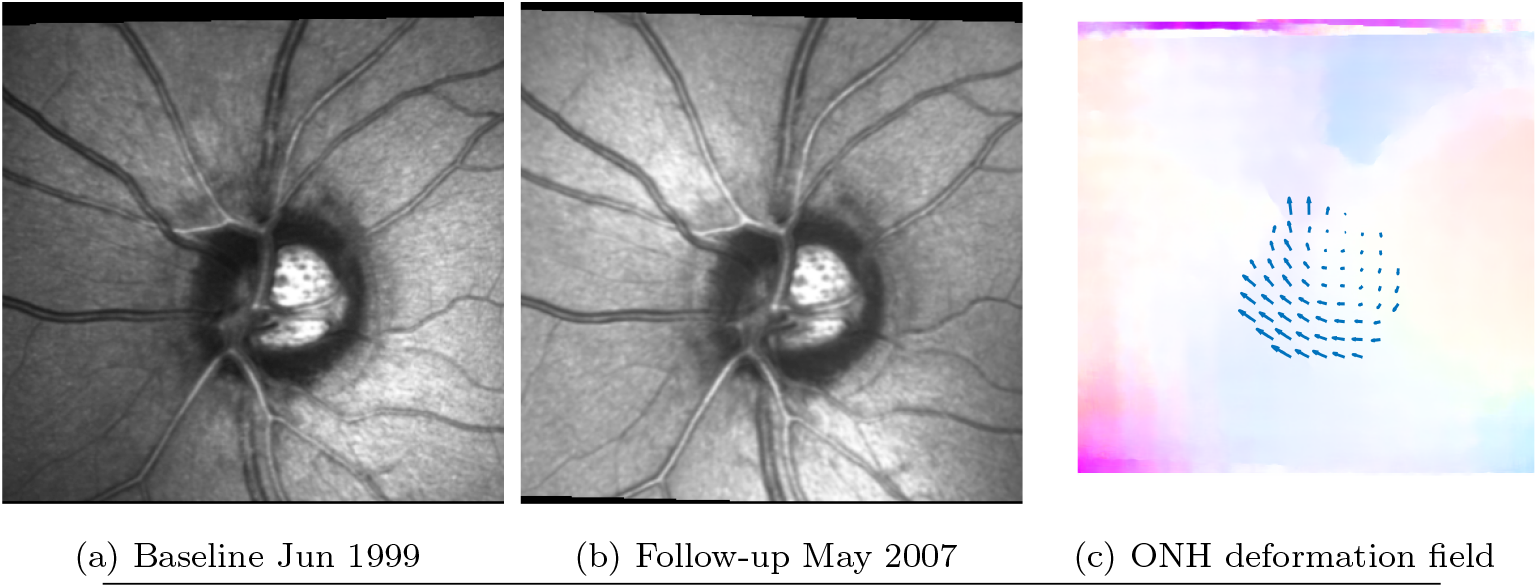
Example of ONH deformation observed in a normal DIGS study eye (human). Deformation field estimated using the DDCNet-Multires method visualized using a color coding scheme shown in Figure 3. For quick inference, deformation field within the optic disk region also shown as arrow plot.

**Figure 7:**
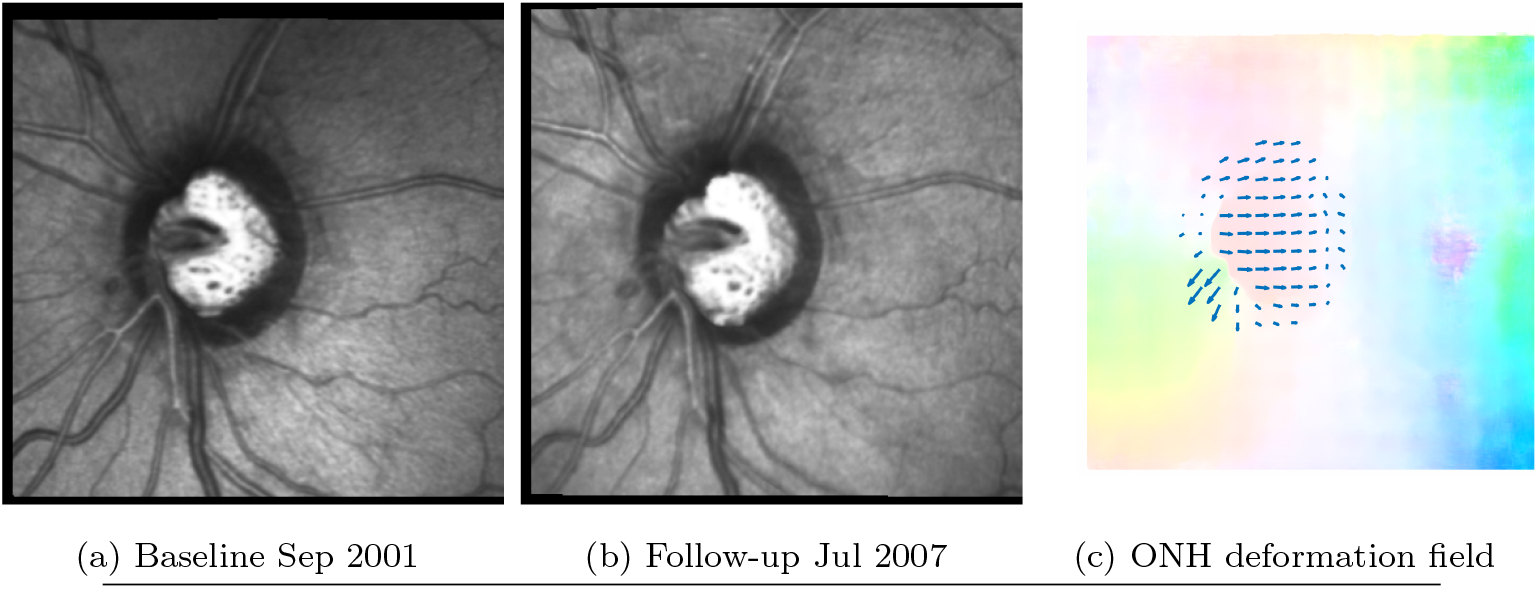
Example of ONH deformation observed in a progressing DIGS study eye (human). Deformation field estimated using the DDCNet-Multires method visualized using a color coding scheme shown in Figure 3. For quick inference, deformation field within the optic disk region also shown as arrow plot.

## DISCUSSION

Though glaucoma detection is established clinically^37^, objective measures or computationally-derived biomarkers of glaucoma progression from imaging and functional measurements are not yet well-defined^38^. *In silico* models of progressive loss of structure (RGC and RGC axons) and function (dysfunction, coma and death of RGCs) in glaucoma have been developed to assess and validate biomarkers of structural and functional progression^39,40^. Structural appearance and functional states of RGCs and nerve fibers may be influenced by ocular conditions (e.g. intraocular pressure fluctuations), systemic conditions (e.g. pulsatile blood flow, eye movements and so forth), testing conditions (e.g. illumination changes, exam quality) and inherent variability of the instruments in capturing the true structural and functional states. Experimental glaucoma studies bridge the gap in accelerating observation of glaucomatous changes in animal models of glaucoma^41–43^.

While various computational methods exist for estimating the motion field that causes structural deformation, it is highly difficult to know the true dense deformation of the structure. This problem is exacerbated for methods estimating ocular deformation as the true dense deformation field is impossible to know even when the ocular images are collected under strict experimental condition. The difficulty here is not with controlling the ocular conditions, but with the continuum nature of the structure and the associated difficulty in knowing its true dense deformation field. The learning-based CNN-architectures were trained using generic image sequences where true dense motion field causing the scene deformation are known. It is common for learning-based optical flow methods to use computer-animated image sequences where the dense deformation field is predetermined or precisely controlled at the pixel and sub-pixel levels. This also significantly alleviates the need for collecting thousands of retinal image sequences under stricter experimental conditions for training the CNN models for estimating the retinal deformation. The performance of these generic optical flow networks for detecting ONH deformation can be further improved by fine-tuning these networks on ONH image sequences from non-human experimental glaucoma studies as well as from clinical studies under controlled conditions.

Digital image correlation methods (DICs) available for estimating structural deformation from digital images^44–46^ is a sub-category of optical flow estimation procedures. Previously, DIC methods have been used to estimate deformation in the cornea, sclera^47^, optic nerve head^48^ and in the lamina cribrosa^14^ from digital image sequences.

For study eyes in the experimental glaucoma study, diagnostic performance of the candidate biomarkers were higher for the classical computational methods (TCA, POD) when compared to the learning-based methods (DDCNet-Multires, FlowNet2, FlowNet-Correlation). For the patient cohort in the DIG study, the diagnostic performance of the learning-based methods were higher for the learning-based methods when compared to the classical computational methods. Upon further inspection, we observed that the lower diagnostic accuracy in the LEG study was primarily driven by large errors in aligning baseline and follow-up exams collected using TopSS confocal system (see Figure 8 for an example). Therefore, the estimated deformation field included both pathologically or experimentally induced ONH deformation as well as errors in aligning ONH topographies. Improving the sequence alignment (using accurate rigid alignment procedures–to avoid introducing artificial deformation) before deformation analysis is likely to improve the overall accuracy of the optical flow methods. No significant image alignment issues were observed in the DIGS study sequences collected using HRT confocal system.

**Figure 8:**
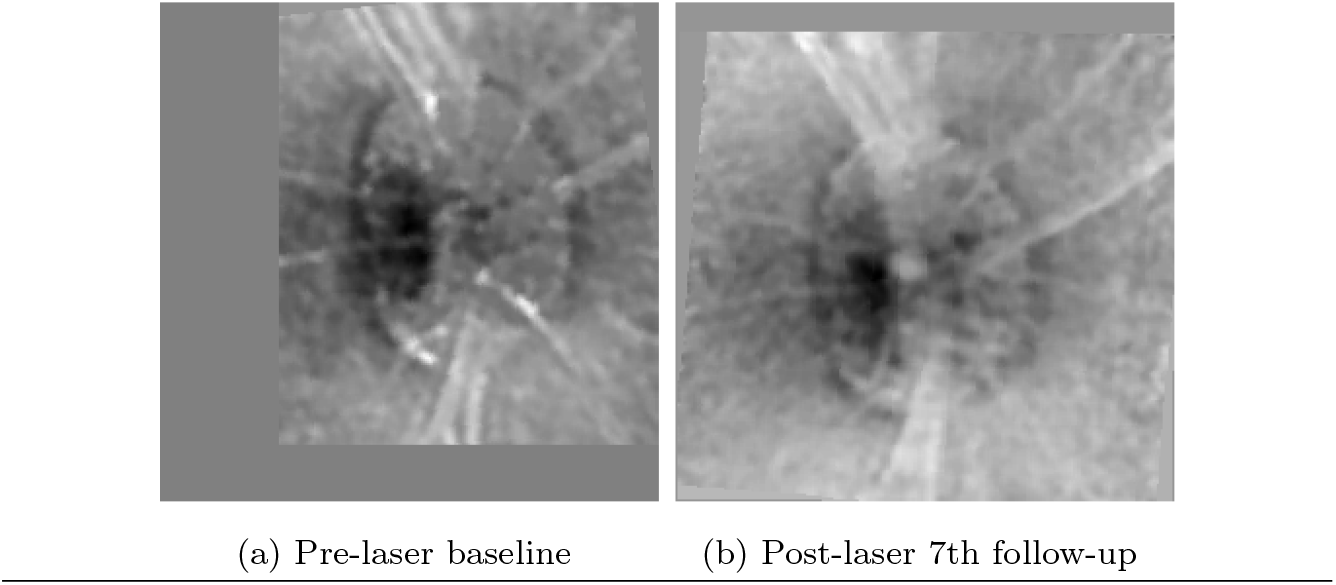
Example of ONH registration error in the LEGS dataset.

Classical POD and TCA methods make an explicit attempt to account for variability in the imaging data due to measurement noise, illumination variability between baseline and follow-up exams as well as systemic sources of variability in the imaging data. Structural biomarker presented in this work is based on estimated deformation of retinal locations. Therefore, incorporating non-pathological sources of imaging variability may further improve the diagnostic performance of the biomarkers based on ONH structural deformation estimated from image sequences.

In this work, we devised a structural biomarker based on the mean ONH deformation magnitudes. Using the estimated dense deformation (i.e. optical flow field), we can easily build other deformation summary statistics such as maximum deformation magnitude, minimum deformation magnitude, median deformation magnitude, and histogram or distributions of deformation magnitude and angles. More importantly, these dense deformation estimates are also useful for estimating underlying mechanical characteristics of the ocular tissues and membranes as elastographic maps comprised of elastic moduli of the constituting elements of the ONH^49^. A detailed description of the elastographic maps as biomarkers of glaucoma progression will be presented in future work.

A typical limitation of learning-based methods is the need for thousands of retinal datasets for training the network to achieve high diagnostic accuracy. One of the advantages of the proposed learned-based method is the reduced requirements for retinal image sequences for training the network for estimating retinal and optic nerve head deformation. The learning-based methods that we investigated in this study were trained using generic image sequences with known dense deformation estimates. Though the networks were not trained using any retinal dataset, all of them were able to estimate ONH deformation from the longitudinal ONH image sequences with moderate to high diagnostic accuracy.

The learning-based methods that we presented in this work estimate ONH deformation from two longitudinal scans of the ONH. The method can be extended to estimate a temporal deformation profile using one baseline and more than two follow-up scans of the ONH. Such an approach is expected to provide a dense trajectory of ONH structural deformation and can be useful for predicting glaucoma progression more accurately.

At present, the learning-based methods presented are based on 2D (either topography or reflectance) scans of the ONH. These methods, framework and design strategies can be extended to estimating volumetric deformation of the ONH using longitudinal 3D scans of the ONH.

## CONCLUSION

Dense ONH structural changes estimated using the optical flow methods can provide insights into the changes in the ONH architecture and associated structural reorganization in glaucoma. Therefore, biomarkers based on raw measures of underlying structural deformation can not only be useful for detecting glaucoma progression but can also be useful for understanding the characteristics of the ONH deformation in patients with varying risk factors such as age, race and other systemic conditions. Our proposed strategies for training the learning-based optical flow methods fully or predominantly using non-ocular image sequences of scenes with known dense deformation field significantly reduces or eliminates the need for collecting training retinal image sequences under stricter experimental conditions. These principles were validated by the higher diagnostic performance of these learning-based methods in both experimentally controlled study eyes and in clinical patient cohorts. Performance of these methods can likely be further improved by fine-tuning these networks using longitudinal ONH sequences collected under stricter experimental conditions. These networks and principles can be extended for estimating volumetric ONH structural changes using 3D optical coherence tomography sequences. The proposed confocal-based structural biomarker of glaucoma progression can be extended to derive similar structural biomarkers from various 2-D scans (e.g. circle scan, *en face* images) and volume scans acquired using optical coherence tomography.

## SOFTWARE

The DDCNet-Multires model along with necessary instructions for running the software are available to the public in the following URL: https://www.computationalocularscience.com/software/

## Data Availability

Code availability: The DDCNet-Multires model along with necessary instructions for running the software are available to the public in the following URL: https://www.computationalocularscience.com/software

## ACKNOWLEDGEMENT

The authors thank Claude F. Burgoyne, M.D., Devers Eye Institute, Portland, OR for providing us with the LEG study topographic library; and Linda Zangwill, Ph.D., University of California San Diego, San Diego, CA for providing us with the clinical DIGS dataset for evaluating the candidate biomarkers of glaucoma progression developed in this work. The contents presented here are solely the responsibility of the authors and do not necessarily represent the official views of the funding agencies.

## Notes

**GRANT SUPPORT:** Research supported in part by an unrestricted startup fund and a Herff graduate fellowship, Herff College of Engineering, The University of Memphis; and a summer student fellowship from Fight for Sight.

### Competing Interest Statement

The authors have declared no competing interest.

### Funding Statement

Research supported in part by an unrestricted start-up fund and a Herff graduate fellowship, Herff College of Engineering, The University of Memphis; and a summer student fellowship from Fight for Sight.

### Author Declarations

The UCSD Institutional Review Board approved the study methodologies and all methods adhered to the Declaration of Helsinki guidelines for research in human subjects and the Health Insurance Portability and Accountability Act (HIPAA).

